# Translation, Validation, and Application of Indonesian Genetic Literacy Questionnaires for Medical Students

**DOI:** 10.64898/2026.04.17.26350524

**Authors:** Rahmat Azhari Kemal, Rahma Dhani, Arya Marganda Simanjuntak, Arimbi Indhira Rafles, Hana Xaviera Triani, Triyanda Miftahur Rahmi, Vinndi Azriel Akbar, Firdaus, Bayu Fajar Pratama, Zulharman

## Abstract

**Background:** Increasing relevance of genetics and molecular biology in medicine necessitates greater genetic literacy among current and future healthcare workers. To assess the literacy level, a validated genetic literacy questionnaire is needed. Therefore, a standardised Indonesian-language genetic literacy questionnaire is essential.

**Aims:** We aimed to translate and validate three genetic literacy questionnaires (PUGGS, iGLAS, and UNC-GKS) for use among Indonesian medical students. We then evaluated genetic literacy levels using one of the validated questionnaires.

**Methods:** The PUGGS, iGLAS, and UNC-GKS questionnaires were translated into Indonesian and then reviewed by an expert panel for translational accuracy and conceptual appropriateness. Back-translation was performed to confirm validity. Initial Indonesian versions of the questionnaires underwent cognitive pre-testing with 12 undergraduate medical students. After refinements, the questionnaires were validated among 34 first-to third-year medical students. The Indonesian version of UNC-GKS questionnaire was then used to assess genetic literacy of 486 medical students comprising 228 preclinical medical students, 187 clerkships, and 71 residents.

**Results:** The Indonesian versions of PUGGS (Cronbach’s α = 0.819) and UNC-GKS (α = 0.809) demonstrated good reliability, while iGLAS showed poor reliability (α = 0.315). Among the 486 students tested, 56% demonstrated moderate overall genetic literacy, and only 15.2% demonstrated good overall literacy. Basic genetic concepts were relatively well-understood with 54.3% having good literacy. On the contrary, gene variant’s effects on health were poorly understood with only 9.7% having good literacy. Inheritance concepts were moderately understood with 24.9% having good literacy.

**Conclusion:** The Indonesian translations of PUGGS and UNC-GKS are reliable tools for assessing genetic literacy among medical students. Using UNC-GKS, we observed predominantly moderate genetic literacy levels. Curriculum improvement to better integrate genetics education is essential to support its clinical applications.

**PRACTICE POINTS:**

- Genetic literacy is needed to fully utilize genetic and genomic application in medicine.
- Adaptation of PUGGS and UNC-GKS genetic literacy questionnaires into Indonesian language resulted in valid and reliable questionnaires to assess genetic literacy among medical students
- Moderate genetic literacy level indicates the need for genetics curriculum update in medical schools in Indonesia, especially in the area of gene variant’s effect on health

## BACKGROUND

Advances in genetics, genomics, and genomic-based technologies are becoming increasingly relevant in all areas and layers of modern society. In the field of medicine, genetics and genomics are applied in diverse areas including prenatal testing, personalized medicines, and even direct-to-consumer genetic testing.^1^ These advances facilitate novel diagnostic capabilities and targeted therapeutic strategies. For instance, genomic technologies can aid in diagnosing patients with rare diseases, congenital anomalies, or developmental disorders. Although such diagnosis may not always lead to immediate treatment options, ending the diagnostic odyssey can offer psychological relief and closure to patients and their families.^2^ In oncology, genetic testing plays a crucial role in informing treatment choices as well as identifying at-risk family members who may benefit from early screening and preventative interventions.^3^ Furthermore, genetics is increasingly integrated into reproductive technologies, such as pre-implantation genetic screening and non-invasive prenatal testing.^4^

To ensure the effective application of genetic and genomic technologies, medical professionals must possess adequate genetic literacy.^5^ Genetic literacy is defined as the sufficient knowledge and understanding of genetic principles that enables individuals to comprehend, interpret, and apply genetic information. It can be measured from the familiarity of genetic terminology, clinical skills, and factual knowledge regarding genes and hereditary traits.^6^ For medical professionals, especially physicians, genetic literacy is essential for identifying potential genetic disorders, delivering appropriate care, and facilitating counselling.^7^ Deficit in this area may lead to misdiagnosis, suboptimal treatments, and unnecessary or inappropriate use of genetic testing.^7^

In the Asia Pacific region, including Indonesia, low levels of knowledge regarding genetics, clinical genetics, and genetic counselling among healthcare workers may present a significant barrier to the advancement of genetic and genomic services in this region.^8^ As medical school is the primary education program to obtain clinical genetic knowledge, assessing genetic literacy among medical students is critical. Several studies revealed concerning patterns in Indonesia. For example, a nationwide survey involving 492 Indonesian medical students found that only 24.59% of participants achieved a sufficient genetic literacy score of above 50%.^9^ In contrast, a similar study involving 1,003 Indonesian medical students reported higher levels of familiarity and genetic literacy, with the average scores of 5.63 ± 0.96 (out of 7) and 6.37 ± 0.83 (out of 8), respectively.^7^ The contrasting results may be attributed to differences in the questionnaires used.

Several questionnaires have been developed to assess genetic literacy, including the Rapid Estimate of Adult Literacy in Genetics (REAL-G)^10^, the Public Understanding and Attitudes towards Genetics and Genomics (PUGGS)^1^, the Genetic Literacy and Attitudes Survey (iGLAS)^11^, and the University of North Carolina Genomic Knowledge Scale (UNC-GKS).^12^ These questionnaires have demonstrated good validity and reliability. However, to date, no standardized or validated genetic literacy assessment tools are available in the Indonesian language. This absence presents a critical gap in the systematic evaluation of genetic literacy in Indonesian medical education.

To address this need, our study aimed to translate and validate genetic literacy questionnaires for precise and culturally appropriate assessment among Indonesian medical students. We subsequently utilized the validated questionnaire to assess the genetic literacy of medical students at various stages of education from a public university.

## METHODS

Three genetic literacy questionnaires used in this study were the International Genetic Literacy and Attitudes Survey (iGLAS)^11^, the Public Understanding and Attitudes towards Genetics and Genomics (PUGGS)^1^, and the University of North Carolina Genomic Knowledge Scale (UNC-GKS).12 From the iGLAS, items used were 20 questions regarding genetic knowledge. From the PUGSS, items used were 16 questions regarding modern genetic & genomic knowledge and 9 questions regarding gene-environment interaction knowledge. From the UNC-GKS, items used were 19 questions regarding genetic knowledge divided into basic concepts, gene-health relationship, and inheritance subsections. Response options in PUGGS and UNC-GKS questionnaires were “true”, “false”, and “don’t know”.

The translation and adaptation process followed the WHO Guidelines on Translation and Adaptation of Instruments.^13^ The process consisted of the following steps:

1. Forward translation. Each questionnaire was translated from English into Indonesian by a research team member proficient in English and knowledgeable in genetics.
2. Expert panel review. The panel consisted of the initial translator, an expert in (medical) genetics and molecular biology, and an expert in instrument development & translation. In this step, the panels reviewed the items to evaluate whether each item adequately addresses core ideas and has good content validity.
3. Back translation. The Indonesian translation was translated back into English by an expert outside of the research team members. The expert had good command of English and expertise in molecular biology. The back-translated questionnaire was compared to the original English version to assess translation validity.
4. Pretesting and cognitive interview. The draft questionnaires were given to 13 medical students from a public university. All students were in their preclinical year, with 3 students in their first year, 3 students in their second year, and 7 students in their third year. The respondents were asked about the following:
  a. What the question/statement was about
  b. Could the respondent paraphrase it in their own words
  c. What the respondent thought when they read the terms used in the question/statement
  d. How the respondent chose their answer
  e. Whether there were any terms that were not understood or inappropriate

Final adapted questionnaires were tested for internal consistency in a separate sample of 34 medical students of a public university. All students were in their preclinical year, with 12 students in their first year, 11 students in their second year, and 11 students in their third year. Cronbach’s alpha was calculated using SPSS software for each questionnaire.

One of the validated questionnaires was used to assess genetic literacy among medical students at an Indonesian public university in 2022. The students were grouped based on educational stage into preclinical students (years 1-3), clerkship students, and resident doctors. First to third year preclinical students were quota-sampled according to the proportion calculated by Slovin’s formula. Clerkship students were sampled from the total population calculated by Slovin’s formula. Due to the smaller population size of residents, total sampling was employed. The students were given an online form of the validated questionnaire. Percentage of correct answers was categorized into good (≥75%), moderate (56-74%), and low (≤55%) genetic literacy. This research has received ethical clearance from Faculty of Medicine Universitas Riau (No: B/105/UN19.5.1.1.8/UEPKK/2022).

## RESULTS

### Translation Process

In this study, three questionnaires, namely PUGGS (item code Pxx), iGLAS (item code iGKxx), and UNC-GKS (item code UNCxx) have been translated from English to Indonesian language. Based on expert panel discussion, there were several suggestions regarding the wording of the statements (Table S1). The experts raised a point on the required background knowledge, suggesting that the students might not be able to answer the questions due to a mismatch between the questions and the coverage or content of genetics in the curriculum.

After rewording, the questionnaires were given to medical students for pretesting. Students reported that they have understood the questions/statements. However, there were several complex sentences, for example the P06 question, that required re-reading for better understanding. The students also proposed rewording and/or restructuring several questions (Table S2). For example, an additional explanation in UNC09 was added for better understanding of “*gejala*” (symptoms). Finally, the students reported difficulty in answering the questions, not because of the language but because of the low familiarity and/or understanding of genetic terms and concepts.

### Validation results

Internal consistency was tested for each questionnaire among 34 respondents, consisting of 11 third-year students, 11 second-year students, and 12 first-year students. The mean score between different study years was not statistically different (p>0.05). Therefore, each questionnaire was analyzed as a single group. Cronbach’s alpha analysis showed that the PUGGS questionnaire demonstrated good reliability (α = 0.819). This questionnaire consists of two sections: modern genetic knowledge (P01 – P16) which had a Cronbach’s alpha of 0.733, and gene-environment interaction knowledge (P17 – P25) that had a Cronbach’s alpha of 0.712, respectively. The UNC-GKS questionnaire also demonstrated good reliability (α = 0.809). However, subsections had varied reliability. Basic concepts (UNC1 – UNC6), gene and health (UNC7 – UNC12), and inheritance (UNC13 – UNC19) had Cronbach’s α of 0.487, 0.781, and 0.506, respectively.

The iGLAS questionnaire had the lowest reliability (α = 0.315). Four items with good validity were regarding DNA base units (iGK02, 70.6% correct answers), the main function of genes (iGK05, 97.1% correct answers), the number of human chromosome pair (iGK08, 73.5% correct answers), and the definition of non-coding DNA (iGK14, 41.2% correct answers). Since almost all respondents answered iGK05 correctly, this question did not correlate significantly with the total iGLAS score. However, iGK02, iGK08, and iGK14 had strong correlation with the total iGLAS score with correlation coefficient of 0.536 (p=0.001), 0.671 (p<0.001), and 0.589 (p<0.001), respectively.

### Inter-questionnaire comparison

While having the least reliability, only the iGLAS score correlated significantly with both the PUGGS (c = 0.369, p<0.05) and the UNC-GKS (c = 0.608, p<0.001) questionnaires. The PUUGS and the UNC-GKS scores did not correlate significantly to each other (p>0.05). Across the three questionnaires, similar questions did not have significant correlation. As an example, the P01, iGK01, and UNC03 are asking about the genome definition. Only 3 (8.8%) respondents answered all three questions correctly (Table S3). Most respondents answered incorrectly in P01 (29.4% correct answers) and iGK01 (32.4% correct answers), but UNC03 was answered correctly by the majority of respondents (88.2% correct answers). The UNC03 statement on genome definition was more general as “all genetic information”. In P01, the item statement was more specific, mentioning that the genome only consisted of protein-coding genes. The misconception is also better observed in the multiple-choice nature of iGK01 where 58.8% of respondents defined the genome as “all genes in DNA” as opposed to the correct answer of “the entire sequence of an individual’s DNA”.

iGK10 and UNC06 pose a similar question regarding estimated gene numbers in human DNA. The iGK10 is a four-choice question while the UNC06 is a true-false question. More than half (18 students, 52.9%) could answer UNC06 correctly. However, among those 18 students, only 5 students could answer iGK10 correctly. Among 16 students who answered UNC06 incorrectly, only one student chose the right answer in iGK10. In iGK10, most (27 students, 79.4%) overestimated the number of human genes. Chi-square test showed no significance (p>0.05) between these two items.

### Genetic literacy of medical students

We used UNC-GKS to measure genetic literacy in sampled medical students. The Indonesian UNC-GKS is available in Table S4. We chose this questionnaire instead of PUGGS due to the fewer number of questions as well as more relevant subsections. We sampled a total of 486 students consisting of 228 preclinical students, 187 clerkship students, and 71 residents (Table 3). The response rate was only calculated for residents due to the initial target of total sampling. We obtained a 64.0% response rate with 71 respondents for a cohort of 111 residents. The response rate was highest from Anesthesiology & Intensive Care (90.9%), followed by Pulmonology & Respiratory Medicine (89.5%) and Surgery (53.8%), while the lowest response rate was from Obstetrics-Gynecology (14.1%).

**Table 3.** Characteristics of respondents.

| Characteristics | Sample (n) | Population (N) | Percentage from population (%) | Percentage from total sample (%) |
| --- | --- | --- | --- | --- |
| **Preclinical** | **228** | **430** | **53.0** | **46.9** |
| First year | 83 | 156 | 53.2 | 17.1 |
| Second year | 67 | 127 | 52.8 | 13.9 |
| Third year | 78 | 147 | 53.1 | 16.0 |
| **Clerkship** | **187** | **319** | **58.6** | **38.5** |
| **Residency\*** | **71** | **111** | **64.0** | **14.6** |
| ANIC | 20 | 22 | 90.9 | 4.1 |
| OBGYN | 10 | 71 | 14.1 | 2.1 |
| PRM | 34 | 38 | 89.5 | 7.0 |
| SURG | 7 | 13 | 53.8 | 1.4 |

\*ANIC: Anesthesiology & Intensive Care; OBGYN: Obstetrics-Gynecology; PRM: Pulmonology & Respiratory Medicine; SURG: Surgery

The overall score indicated a moderate level of genetic literacy (Table 4). Preclinical students had a significantly lower mean score compared to clerkship students (p<0.05), while no significant differences were observed between other groups. The mean score for basic concepts reflected borderline good literacy and was consistent across all study levels. In contrast, the mean score for the gene-health relationship concepts indicated low literacy. However, residents scored significantly higher than preclinical students (p<0.05). Inheritance concepts were moderately understood, with preclinical students scoring significantly lower than clerkship students (p<0.05).

**Table 4.** Genetic literacy score between study level.

| Study level | Genetic literacy score (Mean ± SD) | | | |
| --- | --- | --- | --- | --- |
|  | Total | Basic concepts | Gene & Health concepts | Inheritance concepts |
| Preclinical | 61.06 ± 12.72 | 75.07 ± 16.74 | 48.54 ± 17.36 | 59.77 ± 21.08 |
| Clerkship | 64.11 ± 12.31 | 75.13 ± 16.25 | 52.23 ± 19.15 | 64.86 ± 20.59 |
| Residency | 62.49 ± 13.43 | 72.77 ± 16.72 | 54.69 ± 17.63 | 60.36 ± 24.17 |
| Total | 62.44 ± 12.72 | 74.76 ± 16.53 | 50.86 ± 18.22 | 61.82 ± 21.46 |

Categorisation of scores resulted in a similar conclusion, with most of the respondents (56%) demonstrating moderate overall genetic literacy, and only a small subset (15.2%) exhibiting good literacy (Table 5). This distribution pattern was consistent across study levels, with no significant differences observed (p>0.05). Basic genetic concepts were generally well-understood, with more than half of total respondents (54.3%) having a good literacy in this domain. High levels of understanding were observed on gene composition (UNC1), gene functions (UNC2), the genome (UNC3), and DNA building blocks (UNC5) concepts. However, concepts related to gene stability (UNC4) and gene numbers (UNC6) were less well-understood, as shown by a high percentage of students choosing the “don’t know” option (Figure 1, Table S5). Notably, fewer residents correctly answered UNC4 compared to other study levels (p<0.05).

**Table 5.**
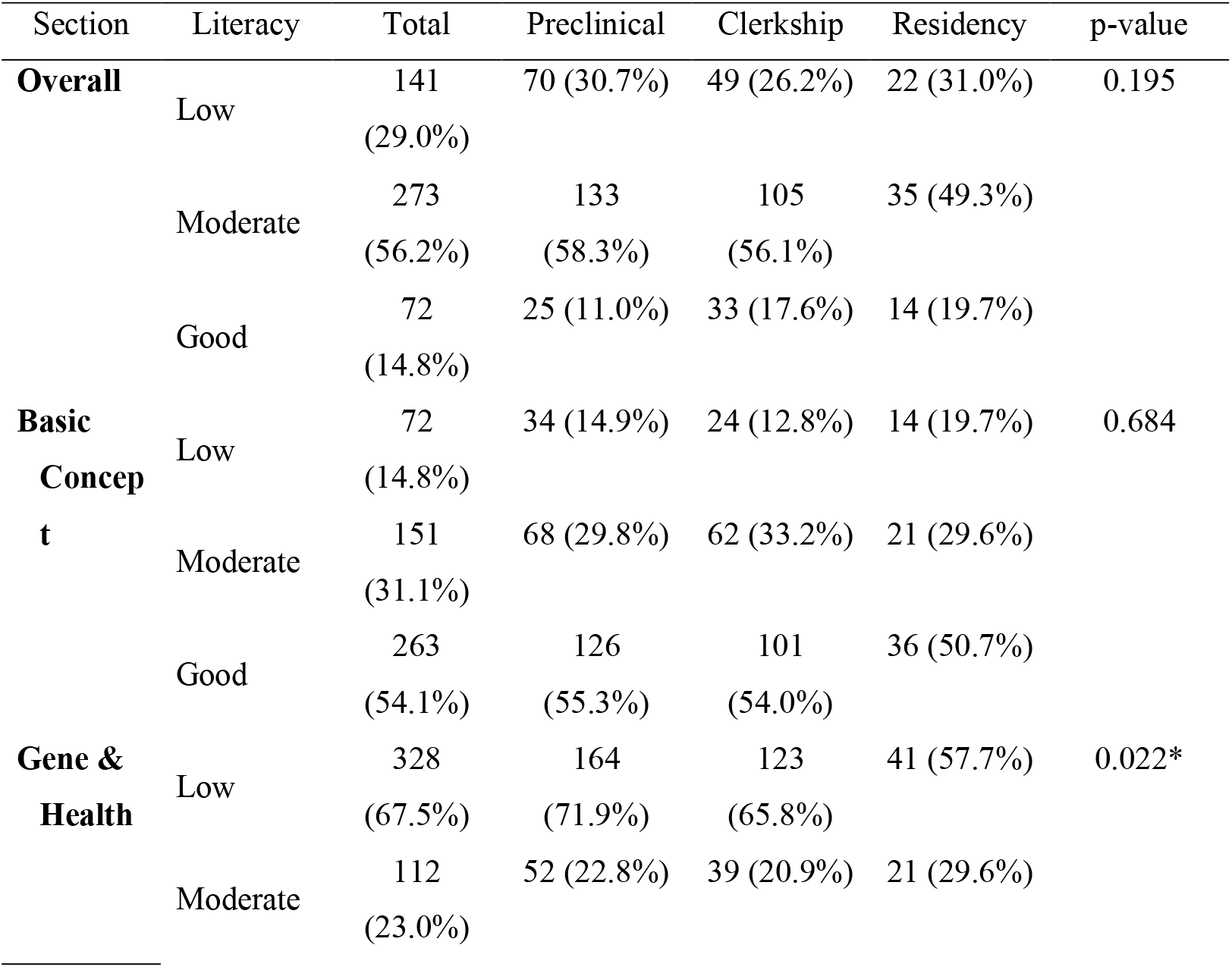
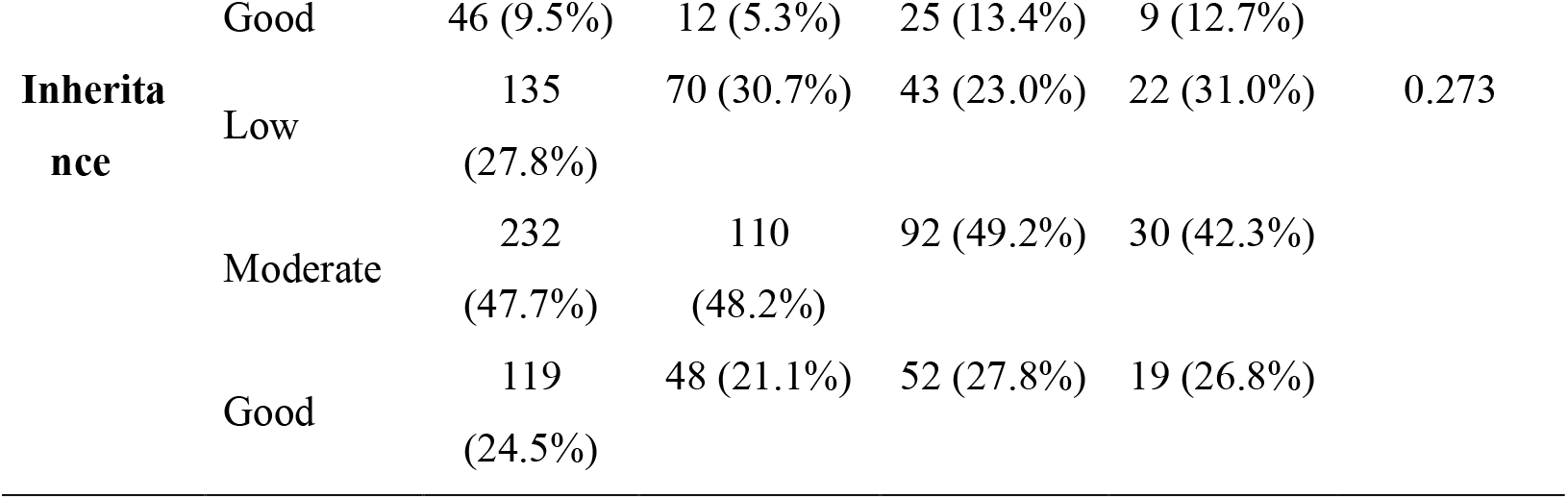
Distribution of genetic literacy category among respondents.

**Figure 1.**
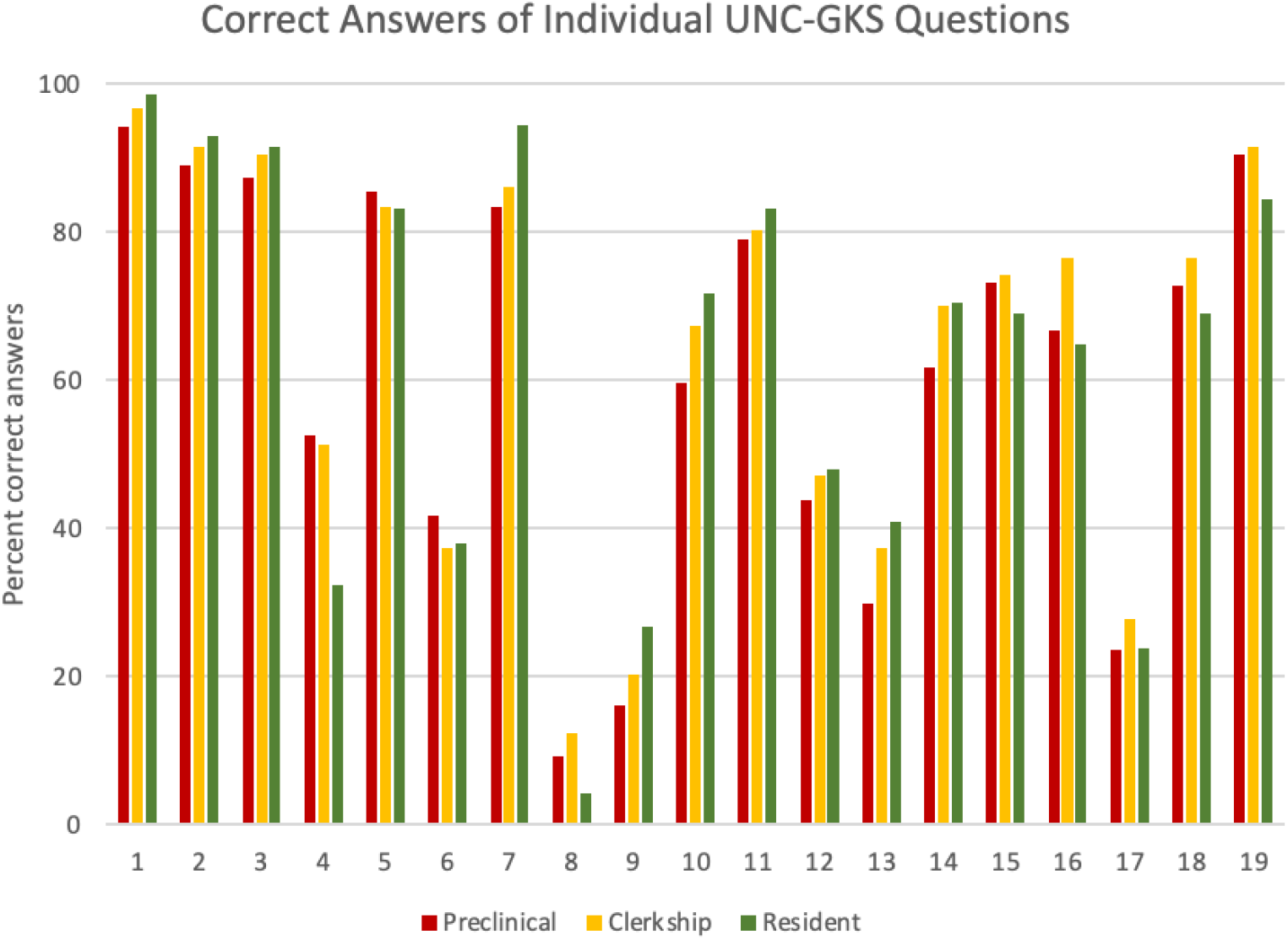
Percentage of correct answers for each individual UNC-GKS showed comparable knowledge between three levels of medical education. Only UNC04 had a significant inter-group difference where fewer residents answered correctly (p<0.01).

Understanding of the effects of gene variants on health was low, with only 9.7% of respondents having good literacy. The distribution of literacy levels was similar across study levels, with the majority of respondents in each subgroup having low literacy and only a minority showing good literacy (Table 5). Preclinical students had significantly lower knowledge compared to clerkship students and residents (p<0.05). Among the gene-health relationship concepts, only the types of variant effects (UNC7) were well-understood, with over 80% of respondents answering correctly (Figure 1, Table S5). Other concepts were moderately or poorly understood. Respondents tended to overestimate the proportion of variants that affect an individual’s health (UNC8), which had the lowest correct response rate across all study levels. Additionally, penetrance effects were also overestimated (UNC9). While respondents recognized that different variants have varying effect sizes (UNC10) and that some variants may reduce the risk of certain disorders (UNC11), more than a third of respondents incorrectly believed that the same genetic variant always manifests the same symptoms (UNC12).

The inheritance concept was moderately understood, with 24.9% of respondents exhibiting good literacy (Table 5). While no significant differences were observed in the literacy category between study levels (p>0.05), preclinical students had a significantly lower mean score compared to clerkship students (p<0.05). Most respondents mistakenly believed that genetic disorders are always inherited (UNC13), indicating insufficient understanding of de novo variants (Figure 1, Table S5). However, respondents correctly recognized that genetic disorders can manifest in only one member of a family (UNC14) and that every individual has a chance for having a child with a genetic disorder (UNC15). Most respondents understood that gene inheritance is not restricted to parent-child pairs of the same sex (UNC16), although the majority incorrectly assumed that physical similarities necessarily correspond to higher genetic similarity (UNC17). The concept of family segregation was partly understood, with majority respondents understanding that if one parent carries a gene variant, their children have a probability of inheriting it (UNC19), but many incorrectly assumed it would be passed down to all children (UNC18).

## DISCUSSION

We successfully translated and validated genetic literacy questionnaires into the Indonesian language. During the translation process, several items were adjusted, and explanatory phrases were added to improve clarity and contextual understanding. This approach is consistent with methodologies used in other validated adaptations of instruments into Indonesian.^14,15^ The final Indonesian version of PUGGS and UNC-GKS questionnaires demonstrated good internal consistency. The reliability of each section of the translated PUGGS questionnaire was comparable to the original instrument. Cronbach’s alpha value for the modern genetic knowledge section was 0.733, compared to the original 0.7. For the gene-environment interaction knowledge section, Cronbach’s alpha value was 0.712, comparable to the original 0.67).^1^ Similarly, the Indonesian version of UNC-GKS also showed strong internal reliability, with a Cronbach’s alpha of 0.809, comparable to the original version’s alpha of 0.86.^12^

We selected the UNC-GKS for further measurement considering its sections being more relevant to the genetic and genomic application in medicine. The basic concepts of genome or gene organization, gene variations, and inheritance are part of the core competencies in medical genetics as outlined by the Association of Professors of Human and Medical Genetics (APHMG).^16^ Notably, the true-false format used in the UNC-GKS may be less effective at revealing misconceptions than the multiple-choice format employed in iGLAS. For instance, most respondents correctly answered a question on human gene numbers in UNC-GKS, but only a minority did so in iGLAS, with most overestimating the number, as previously reported.^17^ However, the inclusion of a “don’t know” response option in the UNC-GKS reduces the likelihood of guessing and enables the identification of specific items associated with high uncertainty among respondents. This feature could highlight specific areas where targeted educational efforts may be required.^12^

During pre-testing, the expert panel expressed concerns that medical students might struggle to answer the questions, not because of linguistic issues but because of insufficient exposure to the genetic content in the medical curriculum. This concern was confirmed during cognitive pre-testing, where panel members reported difficulty understanding the genetic terms and concepts. These challenges were reflected in the final result where the majority of respondents (56%) demonstrated moderate overall genetic literacy, while 28.8% exhibited low literacy (Table 5). This distribution was consistent across all study levels (p-value >0.05). Notably, despite applying a slightly more stringent cut-off for low literacy (≤55%), the proportion of students with low literacy remained lower than in the study by Rujito et al. where 75.41% scored below 50%.^9^ It should be noted that our study was limited to a single public university as compared to a nationwide survey conducted by Rujito et al. Conversely, a national survey by Swandayani et al. found a higher genetic familiarity and literacy among Indonesian medical students.^7^ This difference likely stems from varying instruments. Rujito et al. did not detail their questionnaire,^9^ while Swandayani et al. used an Indonesian REAL-G translation which assesses self-reported familiarity with eight genetic terms.^7^ They proposed that higher scores might reflect Indonesia’s academic system, where medical school applicants, typically natural science majors in high school, may have increased familiarity with scientific terms. This is supported by an observation at an Indonesian public university, where natural science majors demonstrated greater genetic knowledge than social science majors.^18^

The UNC-GKS has three sections addressing different genetic concepts, which are basic genetic concepts, gene-health relationship, and inheritance. Our study revealed that the highest understanding was in the basic genetic concepts compared to gene-health relationship and inheritance concepts. More than half (54.3%) of respondents had good literacy. Our findings aligned with other studies, where similar high confidence in core genetic-genomic concepts but lower in understanding genetics’s contribution to disease was observed.^19^ However, it is important to note that in both the original and Indonesian versions of UNC-GKS, the genome is stated as “all genetic information”. While technically correct, this wording may fail to identify misconceptions about genome composition. Our pre-testing with the Indonesian translation of iGLAS found that more than half of the respondents equated the genome with all genes, rather than all DNA sequences.^17^ Similar misunderstandings regarding the relationship between genes and DNA was also observed among Indonesian undergraduate students.18

Good knowledge on interpreting genetic variants and inheritance patterns is relevant to clinical practice. Surgeons need strong genetic literacy for clinical management, especially in cancer, and to communicate genetic test results and counsel patients. However, studies show surgeons lack knowledge and confidence in genetic inheritance, impacting their comfort with genetic testing and counseling.^20^ This barrier should be addressed to facilitate genetics in routine clinical practice. The physicians’ confidence and awareness of genetic testing can be enhanced by providing additional health education regarding this topic.^21^

Unfortunately, only 9.7% of our respondents had good genetic literacy regarding effects of gene variants on health. Similar to Swandayani et al. (2021)^7^, high self-reported familiarity with “variation” did not translate to correct answers (<10%). Indonesian undergraduate students, even healthcare majors (only 71%), struggled with the healthy-carrier concept, indicating misunderstanding of genetic risk manifestation.^18^ Similar trends exist globally. A UK survey revealed only half of medical students were confident in understanding genetics’ role in diseases, despite high confidence in basic genetic concepts (95%) and inheritance patterns (90%).^19^ Poor knowledge on genetic diseases were also reported in clinical-year medical students in Malaysia.^22^ as well as among practitioners, gynecologists, and pediatricians in the Netherlands.^23^ This knowledge gap presents a need for educational interventions to increase understanding on how gene variants affect health and manifest in disease.

The inheritance concept was moderately understood, with only 24.9% of total respondents having a good literacy level. Similarly, medical students and interns in Saudi Arabia reported lower knowledge on genetic inheritance.^24^ In contrast, 90% of UK medical students reported good confidence in understanding inheritance patterns.^19^ Among primary care physicians and medical specialists, overall knowledge on hereditary breast cancer was good. However, knowledge on breast cancer inheritance was more limited.^25,26^ This limited understanding on how genes are inherited in families might be related to limited comprehension of Mendelian concepts. It has been argued that a student’s understanding of Mendelian concepts is essential to support understanding of other inheritance topics.^27^ Supporting this, a small study involving 25 Indonesian pre-service biology teachers reported lower Mendelian inheritance scores compared to other genetic topics.^28^ In a different Indonesian university, many undergraduate students have difficulties in understanding the Mendelian principles and reconstructing chromosomes based on Mendel’s laws.^29^ These difficulties might have arisen since high school. Review of Indonesian high school biology textbooks revealed incomplete coverage of genetic concepts and the presence of potential misconceptions.^30^

The low genetic literacy indicates an urgency to develop genetic learning materials that are relevant in medical sciences and in accordance with the Indonesian Medical Doctor Competency Standard (*Standar Kompetensi Dokter Indonesia*, SKDI). The proportion of genetics content in the current Indonesian medical curriculum is minimal.^9^ It has been argued that the current curriculum does not equip the future doctors with the basics of genomics and genetic screening to support the Indonesian health system transformation plan in adopting genetic and genomic screening.^31^ The Indonesian Medical Council and relevant authorities in Indonesian medical education can collaborate with the Indonesian Society of Human Genetics to draft core genetic knowledge and competencies for medical doctors. Such an approach can be adapted from the Association of Professors of Human and Medical Genetics (APHMG) in developing consensus core competencies.^16^ Consensus for definition and measurement of genetic literacy is also needed.^32,33^ For the current physicians and specialists, online-based continuing medical education programs in genetic or genomic medicine can be developed.^19^ Online training was shown to help surgical oncologists and nurses to have a positive attitude, confidence, and self-efficacy to provide pre-test genetic counselling to breast cancer patients.^34^ New teaching resources can also be developed. We have developed a genetic module based on a Learning Management System that can be used for self-paced study.^35^ The Indonesian version of UNC-GKS can be utilised to evaluate the new curriculum as well as evaluate the educational interventions.

While our study only assessed genetic literacy among 486 medical students in a single institution, the result reflected similar studies with larger and broader samples.^7,9^ We hope the adoption of the Indonesian version of UNC-GKS for genetic literacy measurement in Indonesia will provide a more representative figure. The questionnaire can also serve as a foundational option in determining consensus definition and measurement of genetic literacy in Indonesia.^33,36^

This questionnaire demonstrates its relevance in assessing genetic literacy among medical students. Further studies are needed for other healthcare worker populations such as nurse, pharmacists, and nutritionists, as there is potential for genetic knowledge gaps, as found in a pilot study of Indonesian nursing students.^37^ Genetic literacy is a relevant competency for nurses to provide support and guidance to individuals and families undergoing genetic testing.^38,39^ Additionally, as pharmacogenetics and nutrigenetics are becoming more relevant in medical practice, good genetic literacy is also needed by pharmacists and nutritionists.^40,41^

## CONCLUSIONS

We translated and validated the PUGGS and UNC-GKS genetic literacy questionnaires into Indonesian. Using UNC-GKS, we found moderate genetic literacy among medical students at a public university, with good basic concept understanding but limited knowledge of variant effects and inheritance. This highlights the need to revise Indonesia’s medical genetics curriculum. The validated Indonesian UNC-GKS can evaluate the new curriculum and track improvements. Further validation in other healthcare professionals is recommended. Improved genetic literacy among Indonesian medical and healthcare workers will facilitate genetic technology adoption, leading to better patient care and health outcomes.

## Data Availability

All data produced in the present study are available upon reasonable request to the authors.

## ACKNOWLEDGEMENT

We thank the experts and students contributing to this study. The funding for this study was provided by Universitas Riau research grant *Penelitian Dosen Muda* scheme (No. 1419/UN19.5.1.3/PT.01.3/2022).

## COMPETING INTEREST

The authors declare that there are no competing interests related to the study

## LIST OF ABBREVIATIONS

iGLAS: International Genetic Literacy and Attitudes Survey
PUGGS: Public Understanding and Attitudes towards Genetics and Genomics
UNC-GKS: University of North Carolina Genomic Knowledge Scale

## AUTHORS’ CONTRIBUTION

RAK contributed to research conception and design, data acquisition and analysis, data interpretation, and manuscript writing. RD and AMS contributed to data interpretation and manuscript writing. AIR, HXT, TMR, and VAA contributed to data acquisition and analysis. F and BFP contributed to research conception and design. Z contributed to research conception and design, data acquisition and analysis, data interpretation, and manuscript writing. All authors gave final approval and agreed to be accountable for all aspects of the work.

## Supplementary materials

Please contact the corresponding author to request access to supplementary materials containing the suggestions from expert panel on initial translation (Table S1), suggestions from pretesting and cognitive interview (Table S2), and the final Indonesian version of UNC-GKS questionnaire (Table S4). Other supplementary tables are available below.

**Table S3.** Percentage of correct answer for questions regarding genome definition.

| UNC3 | P01 | iG01 |  |
| --- | --- | --- | --- |
|  |  | Incorrect | Correct |
| Incorrect | Incorrect | 2 (5.9%) | 0 |
|  | Correct | 1 (2.9%) | 1 (2.9%) |
| Correct | Incorrect | 15 (44.1%) | 7 (20.6%) |
|  | Correct | 5 (14.7%) | 3 (8.8%) |

**Table S5.**
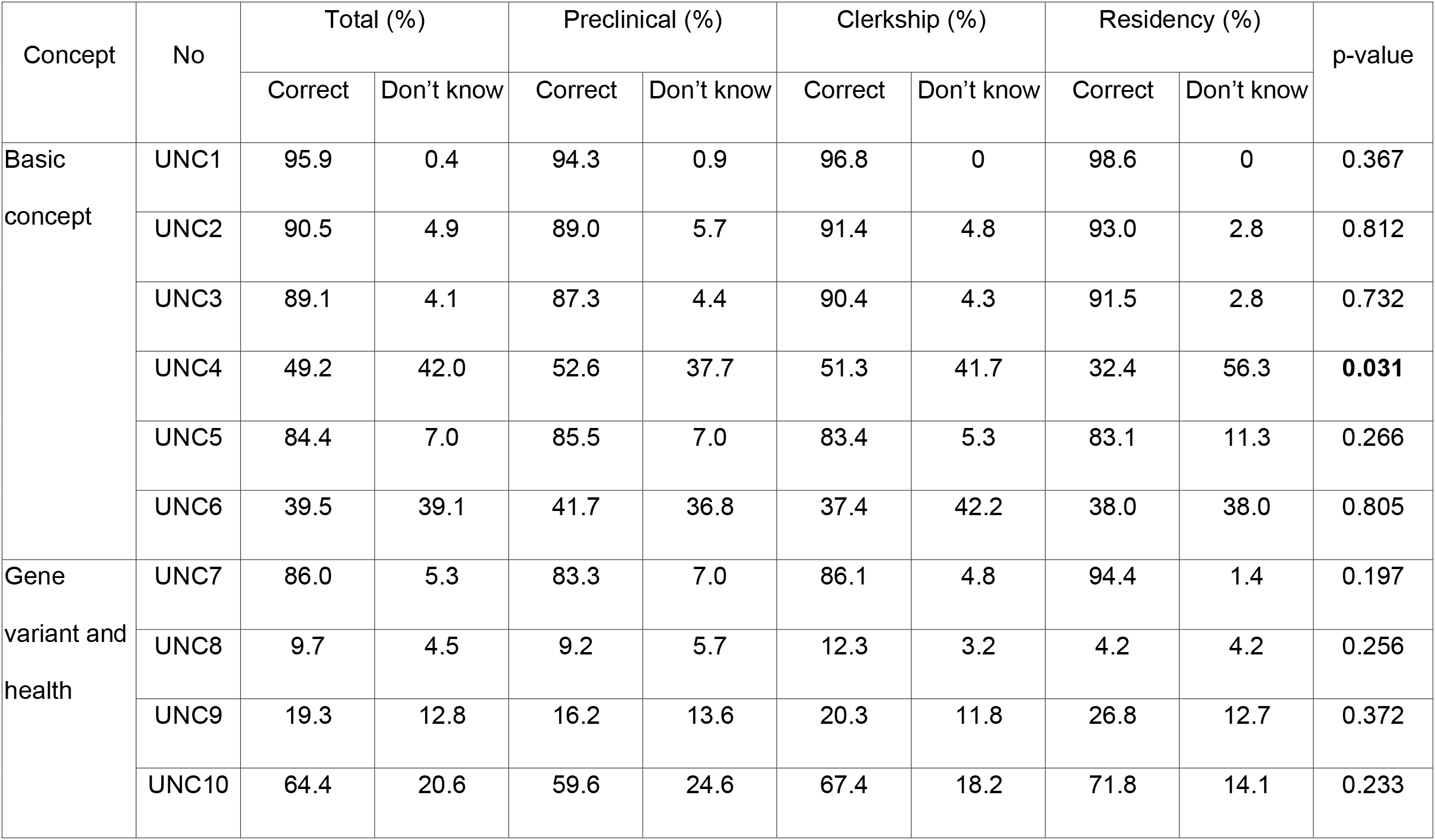
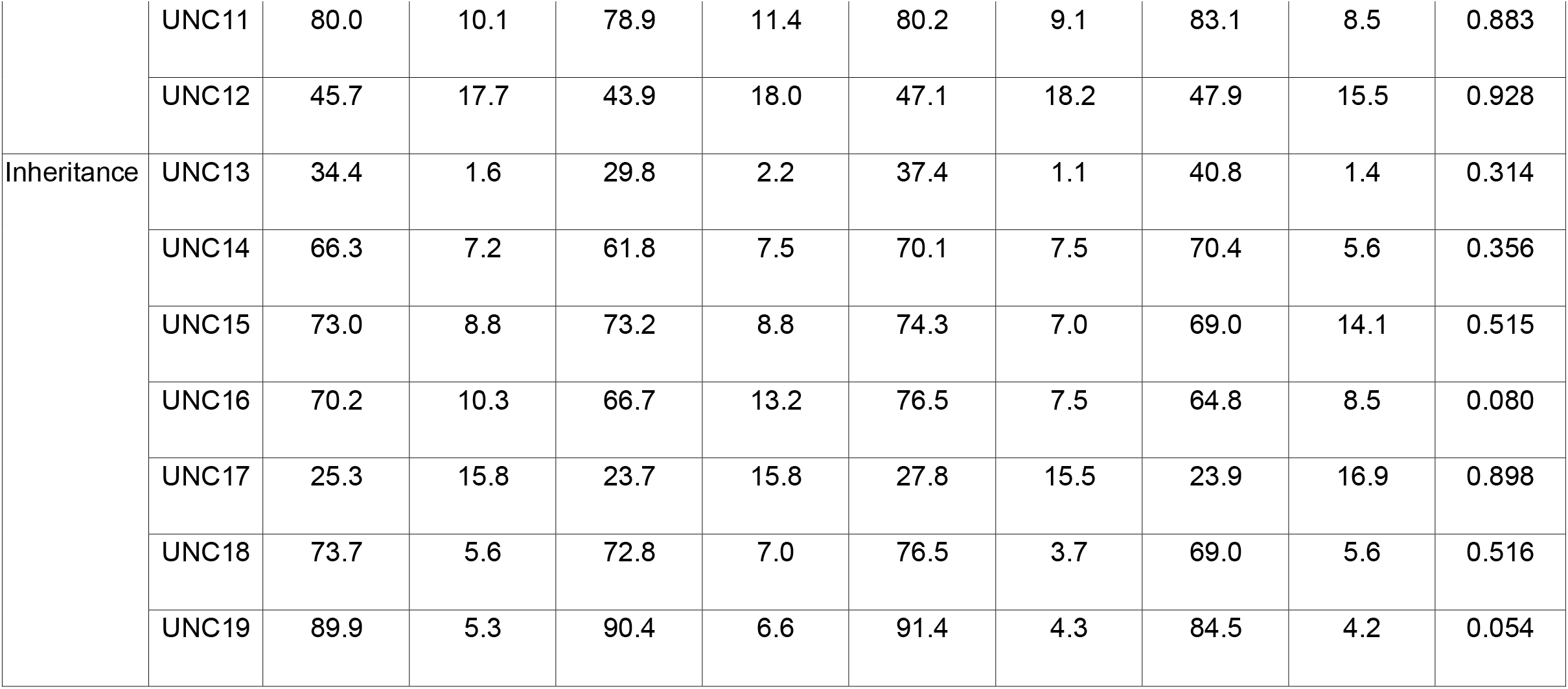
Percentage of correct answer for each UNC-GKS question in different medical study level.

## Notes

### Competing Interest Statement

The authors have declared no competing interest.

### Funding Statement

This study was funded by Universitas Riau (No. 1419/UN19.5.1.3/PT.01.3/2022).

### Author Declarations

Ethical Review Board for Medicine & Health Research of Fakultas Kedokteran Universitas Riau gave ethical approval for this work (No: B/105/UN19.5.1.1.8/UEPKK/2022).

